# Nucleus basalis of Meynert integrity is associated with cognitive dysfunction in Parkinson’s disease independently of locus coeruleus degeneration

**DOI:** 10.64898/2026.07.21.26358398

**Authors:** Alexander Bailey, Etienne St-Onge, Victoria Madge, Neda Shafiee, Jean-François Gagnon, Alain Dagher, D. Louis Collins, Madeleine Sharp

## Abstract

**Background:** Degeneration in the cholinergic nucleus basalis of Meynert (nbM) is thought to contribute to early cognitive deficits in Parkinson’s disease (PD). However, it is unknown whether this relationship is confounded by the parallel degeneration of the substantia nigra (SN) and the locus coeruleus (LC), and whether this relationship is unique to PD.

**Methods:** We conducted a cross-sectional analysis in 112 PD patients (disease duration <10 years) and 46 controls who underwent standard neuropsychological testing, diffusion-weighted and neuromelanin magnetic resonance imaging. Mean diffusivity (MD) of the nbM and neuromelanin signal of the SN and LC were used as proxy measures of neurodegeneration.

**Results:** nbM MD did not differ between PD patients and controls (β=0.05, 95% CI [−0.27, 0.37], *p*=.771), a finding that was replicated using other MRI metrics. Higher nbM MD in PD patients was associated with worse executive function (β=−0.22, 95% CI [−0.39, −0.05], *p*_FDR_=.027), controlling for degeneration in the SN and LC. An association with attention did not survive multiple comparisons correction (β=−0.22, 95% CI [−0.41, −0.02], *p*_FDR_=.112), and there was no association with memory (β=0.08, 95% CI [−0.15, 0.30], *p*_FDR_=.572). When considering both groups jointly, the relationship between increased nbM MD and worse executive function was stronger in PD than controls (β_nbM_*_Group_=−0.30, 95% CI [−0.54, −0.06], *p*_FDR_=.036).

**Conclusion:** Our findings suggest that in early-stage PD, nbM microstructural changes may account for unique variance in executive dysfunction in PD, independent of the effects of LC degeneration, and with a stronger association in PD than controls.

**KEY MESSAGES:** *What is already known on this topic:* Although cognitive deficits may stem, at least in part, from degeneration of cholinergic neurons in the nucleus basalis of Meynert (nbM), it is unknown whether this relationship is confounded by the parallel degeneration of the substantia nigra (SN) and the locus coeruleus (LC), and whether this relationship is unique to PD.

*What this study adds:* The novelty of our approach is that we considered the effects of the nbM, SN, and LC jointly rather than in isolation. We found that in early-stage PD, loss of nbM microstructural integrity was associated with executive dysfunction in PD, independent of the association of LC and executive dysfunction.

*How this study might affect research, practice, or policy:* These findings have important clinical implications because they suggest that the effects of degeneration in the cholinergic and noradrenergic system are at least partially independent and therefore could be targeted separately for remediating cognitive impairments.

## INTRODUCTION

Cognitive deficits are highly prevalent in Parkinson’s disease (PD), even in the early stages of the disease^1^. Findings have suggested that these deficits may stem, at least in part, from degeneration of cholinergic neurons in the nucleus basalis of Meynert (nbM), a basal forebrain nucleus that provides the primary source of cholinergic projections to the cerebral cortex^2^. However, two potential confounders to this relationship remain unaddressed. First, early degeneration in PD also occurs in other subcortical grey matter nuclei that modulate cognition, most notably the dopaminergic substantia nigra (SN) and the noradrenergic locus coeruleus (LC)^3–5^. Second, basal forebrain atrophy is also a common feature of aging, possibly reflecting early Alzheimer’s disease-associated degeneration^2,6^. As such, the degree to which the relationship between nbM degeneration and cognition reflects a unique effect of PD neurodegeneration, and whether this relationship is selective to the nbM, remains unclear.

Research findings from animal and human neuroimaging studies provide converging evidence that integrity of the cholinergic basal forebrain, including the nbM and its cortical projections, supports attentional and working memory processes^7–9^. Studies using structural and diffusion-weighted MRI to quantify atrophy and loss of microstructural integrity in the nbM in PD have shown a relationship between these proxy measures of degeneration and cognitive deficits on measures of executive function and global cognition^10,11^. However, a similar relationship has also been demonstrated in Alzheimer’s disease and in cohorts of older adults without neurodegenerative disease^8,12,13^. Indeed, studies examining whether loss of nbM volume and microstructural integrity in the nbM is greater in PD patients than older adults have yielded mixed results^14–16^. This raises questions about the degree to which cognitive dysfunction in PD is due to PD-specific neurodegeneration or to the effect of age-related processes that are also known to occur in PD.

Degeneration of the LC and the SN is also a prominent feature of early PD, likely precedes nbM degeneration, and contributes to cognitive dysfunction^3,17,18^. In particular, degeneration of the LC in PD, as measured by neuromelanin MRI, has also been associated with impaired executive function^3,18,19^, raising questions about the specificity of these brain-behaviour relationships.

To address these questions, we conducted a cross-sectional analysis of a sample of PD patients without dementia who had a disease duration of < 10 years, and older adults controls who underwent standard neuropsychological testing and neuroimaging. Diffusion-weighted magnetic resonance imaging (DWI) was used to measure mean diffusivity in the nbM, with increased diffusivity used as a proxy measure for loss of microstructural integrity^14^. Neuromelanin MRI was used to measure the bright signal in the SN and LC, with reduced signal used as a proxy measure for degeneration^18^. We hypothesized that loss of microstructural integrity in the nbM in PD patients would be associated with worse cognitive performance, controlling for signal loss in the SN and LC, and that the strength of the relationship would be greater in patients than in controls. We focused on the cognitive domains of attention, executive function, and memory, as these three domains represent cognitive processes thought to be modulated by the cholinergic system^7,11^.

## METHODS

### Participants

PD patients and controls were recruited from the Quebec Parkinson Network (QPN), a registry consisting of PD patients referred by their treating neurologist and older adults interested in participating in research^20^. Participants included in the current study were selected from a larger cohort of participants if they had a Montreal Cognitive Assessment (MoCA) score > 18; PD patients were additionally required to have a disease duration < 10 years^21^. Sample characteristics of the 112 PD patients and 46 controls included in the analysis are presented in **Table 1**. PD patients were tested in their usual ‘On’ dopaminergic medication state. All participants provided informed consent. The study was approved by the McGill University Health Centre Research Ethics Board.

**Table 1.** Sample Characteristics.

| Characteristic | Controls<br>(n = 46) | PD<br>(n = 112) | *p* value |
| --- | --- | --- | --- |
| Males/Females | 11/35 | 76/36 | **<.001** |
| Age | 63.15 (11.51) | 63.30 (9.34) | .937 |
| Education (years) | 15.24 (3.96) | 15.14 (2.7) | .880 |
| Disease duration (years) | - | 3.95 (2.48) | - |
| Hoehn & Yahr stage | - | 1.97 (0.84) | - |
| UPDRS III | - | 27.74 (13.04) | - |
| MoCA | 27.40 (2.99) | 25.47 (3.14) | **.006** |

*Note.* Statistical comparisons conducted using Welch's t-tests or chi-square, with significant *p*-values listed in bold. Values represent mean (standard deviation). UPDRS III, Unified Parkinson's Disease Rating Scale Part III; MoCA, Montreal Cognitive Assessment. Please note that disease duration (in years) was unavailable in 4 PD participants, Hoehn & Yahr stage for 34, and UPDRS III scores for 6 PD participants.

### Clinical and Neuropsychological Assessments

The present study focused on neuropsychological testing of the domains of attention, executive function and memory, as performance in these domains has been found to be correlated with cholinergic function in PD^7,11^. Testing followed Movement Disorder Society recommendations^22^. To address issues of missing data and to ensure consistent domain score definitions across participants, we included only tests with complete data. Attention was assessed with the Trail Making Test Part A (TMT-A, total seconds). Executive function was assessed with the Brixton Spatial Anticipation Test (BSAT; total number of errors) and the Trail Making Test (difference between Part B and Part A, in seconds). Memory was assessed with the Hopkins Verbal Learning Test-Revised (HVLT-R; trials 1-3, total correct) and HVLT-R Trial 4 (delayed recall, total correct). We reverse coded performance on the TMT and the BSAT, such that higher values reflected better performance in all tests. Test scores were standardized within the PD group for PD-only analyses and across the full sample for between-group analyses. Composite scores were computed for executive function and memory and consisted of the mean of the test-specific z-scores. Attention performance was taken as the z-score of TMT-A performance.

### Neuroimaging

The MRI protocol included a 3D T1-weighted magnetization-prepared rapid acquisition gradient echo sequence, a 2D T1-weighted fast spin echo (neuromelanin MRI) sequence, and 2D pulsed gradient spin EPI sequences for DWI, acquired on a Siemens PrismaFit 3T system with a 32-channel head coil. Imaging parameters for the T1-weighted sequences have been previously reported and are provided in **Supplemental Table 1**^18^. DWI imaging parameters included: repetition time (TR) of 6900 ms; echo time (TE) of 64 ms; slice thickness of 2.0 mm for 70 slices; field of view (FOV) of 256 mm; echo spacing of 0.57 ms; EPI factor of 128; 30 diffusion-weighted directions (b-value of 1000); 3 b0 images; 3 reverse encoded b0 images; output matrix size of 256 x 256 x 140 mm; and a voxel size of 2.0 x 2.0 x 2.0 mm^3^. All raw T1 images underwent skull stripping, non-local means denoising, non-uniformity correction, and intensity normalization using the NeuroImaging & Surgical Technologies Longitudinal processing pipeline^23^. T1 images were registered to stereotaxic space using the PD126 template as the registration target^24^.

#### DWI processing and nbM measurements

To create the nbM mask, a probabilistic atlas was used, applying a 50% threshold on the average template to define the final mask^25^. Eddy currents and topup corrections were applied to all DWI images using the FMRIB Software Library^26^. DWI images were visually assessed before and after those corrections. DWI images were denoised via *MRtrix*, corrected for intensity nonuniformity with N4 and aligned to T1 images using *ANTS* SyN^27,28^. *DIPY* was used to estimate the diffusion tensor imaging model and recover diffusivity measures, namely, mean diffusivity, axial diffusivity and fractional anisotropy^29^. All diffusion measures were extracted in native space with region of interest (ROI) masks from the atlas, aligned via non-linear registration to the DWI space of each participant. NbM voxels with significant CSF partial volume (CSF >20%) were excluded from the DWI analysis^30^. Participant T1-weighted scans were up-sampled to a 0.5 x 0.5 x 0.5mm resolution and then non-linearly registered to an unbiased average template. Deformation-based morphometry analysis was conducted to assess local nbM volume differences at the voxel level by computing Jacobian determinants of the deformation fields generated during non-linear registration to the template. Voxel-wise volume difference values were then averaged within the nbM region of interest to derive a single measure of nbM volume difference for each participant relative to the template. Our primary measure of nbM microstructural integrity was mean diffusivity (MD), as MD has been found to be a robust measure of nbM integrity in PD^14^. Exploratory analyses were conducted using axial diffusivity (AD), fractional anisotropy (FA), and nbM volume. Right and left values were averaged.

#### Neuromelanin MRI processing and LC and SN signal extraction

Raw neuromelanin images underwent slice-by-slice normalization to remove any inter-packet or edge-slice intensity differences from the images using the MINC Toolkit^31^. Images were then co-registered to their native T1w image and brought to the PD126 stereotaxic space using the concatenated co-registration warp with the T1w stereotaxic warp. Neuromelanin signal extraction was performed in PD126 stereotaxic space. The SN region of interest was manually defined via expert manual segmentations from a radiologist on an in-house neuromelanin template, while the LC region of interest was defined using a conservative threshold from a probabilistic atlas of brainstem nuclei^32^. Left and right labels were made symmetric to remove any template atlas-related bias to ensure left-right differences found could be attributed to true signal differences. Neuromelanin scores for the LC and SN were calculated as contrast ratios, using the pontine tegmentum and cerebral peduncles as background regions for intensity normalization, respectively, as previously described^18^. The mean of the right and left scores was used in analyses.

### Analysis

All statistical analyses were carried out in R version 4.5.3. Group differences in demographic characteristics were evaluated using Welch’s *t*-tests or chi-square tests. Group differences in MRI-derived measures of neurodegeneration were evaluated using linear regressions with group as a predictor, controlling for age, sex, and education. Associations between nbM DWI measures and performance in each cognitive domain were evaluated using linear regressions on performance controlling for degeneration in the SN and LC (neuromelanin scores), as well as age, sex, and education. These regression models were also conducted in the whole sample with the addition of group as a predictor and its interaction with nbM MD, to determine whether the relationship between nbM integrity and cognition differed between groups. MRI-derived measures of the nbM, SN, and LC were standardized to enable comparison of effect sizes.

Standardization was performed within the PD group for PD-only analyses and across the full sample for analyses comparing groups. Regression models examining the relationship between MRI-derived measures of neurodegeneration and cognitive performance were corrected for multiple comparisons for the three cognitive outcomes using the Benjamini-Hochberg false discovery rate (FDR) procedure (α = .05).

## RESULTS

### Group differences in MRI-derived measures of nbM, SN, and LC integrity

Average nbM MD did not differ between PD and controls (β_Group_ = 0.05, 95% CI [−0.27, 0.37], *p* = .771; **Figure 1**). Older age was associated with higher nbM MD (β_Age_ = 0.06, 95% CI [0.05, 0.07], *p* < .001, **Table 2**). Similarly, average AD (β_Group_= −0.01, 95% CI [−0.34, 0.32], *p* = .954), FA (β_Group_ = −0.22, 95% CI [−0.60, 0.17], *p* = .267), and nbM volume (β_Group_= −0.164, 95% CI [−0.53, 0.20], *p* = .372) did not differ between groups (**Supplemental Figure 1**). As expected, neuromelanin scores were lower in PD patients than controls in both the SN (β_Group_= - 0.77, 95% CI [−1.13, −0.43], *p* < .001) and LC (β_Group_= −0.62, 95% CI[-0.98, −0.26], *p* = .001; **Supplemental Figure 2).**

**Figure 1.**
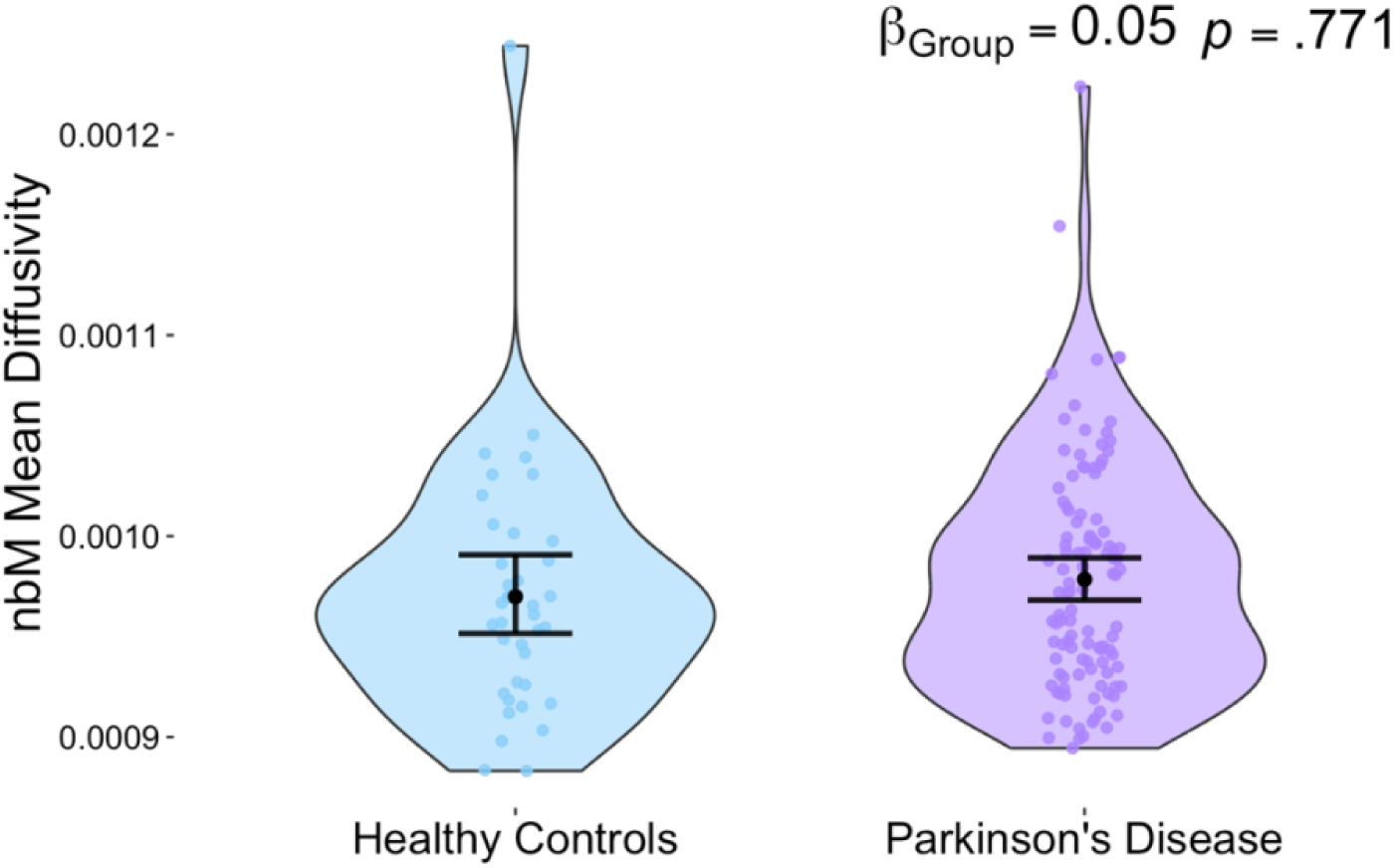
Comparison of nbM MD across groups. Violin plots of nbM MD (average of right and left) in Controls and Parkinson’s patients. The beta estimate and p-value for the group difference were extracted from the linear regression model adjusted for age, sex, and education. Error bars reflect 95% bootstrap confidence intervals.

**Table 2.** Adjusted linear regression model to examine group differences in nbM MD.

| Predictors | $\beta$ | CI (95%) | p value |
| --- | --- | --- | --- |
| Intercept | -3.819 | -4.94, -2.70 | **<0.001** |
| Group | 0.048 | -0.27, 0.37 | 0.771 |
| Age | 0.058 | 0.05, 0.07 | **<0.001** |
| Sex | 0.288 | 0.001, 0.58 | **0.049** |
| Education | -0.003 | -0.05, 0.04 | 0.889 |
| R<sup>2</sup> / R<sup>2</sup> adjusted |  |  | 0.371 / 0.353 |

*Note.* Group refers to PD vs. Controls (0 = Controls; 1 = PD); Sex refers to female vs. male (0 = Female; 1 = Male); Education was measured in years.

### Group differences in cognitive performance

PD patients performed significantly worse than controls in attention (β_Group_= −0.51, 95% CI [−0.88, −0.13], *p* = .008), executive function (β_Group_= −0.47, 95% CI [−0.77, −0.18], *p* = .002), and memory (β_Group_= −0.74, 95% CI [−1.07, −0.40], *p* < .001; **Supplemental Figure 3)**. Performance on the individual tests used to compute the composite scores is presented in **Supplemental Table 2.**

### Relationship between nbM integrity and cognitive performance in Parkinson’s disease

In linear regression models adjusted for SN and LC neuromelanin, age, sex, and education, higher nbM MD was associated with worse attention (β_nbM_ = −0.217, 95% CI [−0.41, −0.02], *p* = .032) and worse executive function (β_nbM_= −0.219, 95% CI [−0.39, −0.05], *p* = .012), but not with memory (β_nbM_= 0.078, 95% CI [−0.15, 0.30], *p* = .491; **Supplemental Table 3**). After FDR correction, however, only the relationship between nbM MD and executive function remained significant (β_nbM_ = −0.219, 95% CI [−0.39, −0.05], *p* = .027; **Table 3**). Lower LC neuromelanin signal was associated with both worse attention (β_LC_= 0.444, 95% CI [0.27 – 0.61], *p* < .001) and executive function (β_LC_= 0.252, 95% CI [0.11, 0.40], *p* = .001), but not memory. These relationships persisted after FDR correction (**Table 3**). SN neuromelanin signal was not associated with performance in any cognitive domain (**Table 3**). Similar associations with executive function were observed for nbM FA and volume. nbM volume was also associated with memory performance. When nbM AD was used, however, no statistically significant relationships to cognition were found (**Supplemental Table 4)**.

**Table 3.**
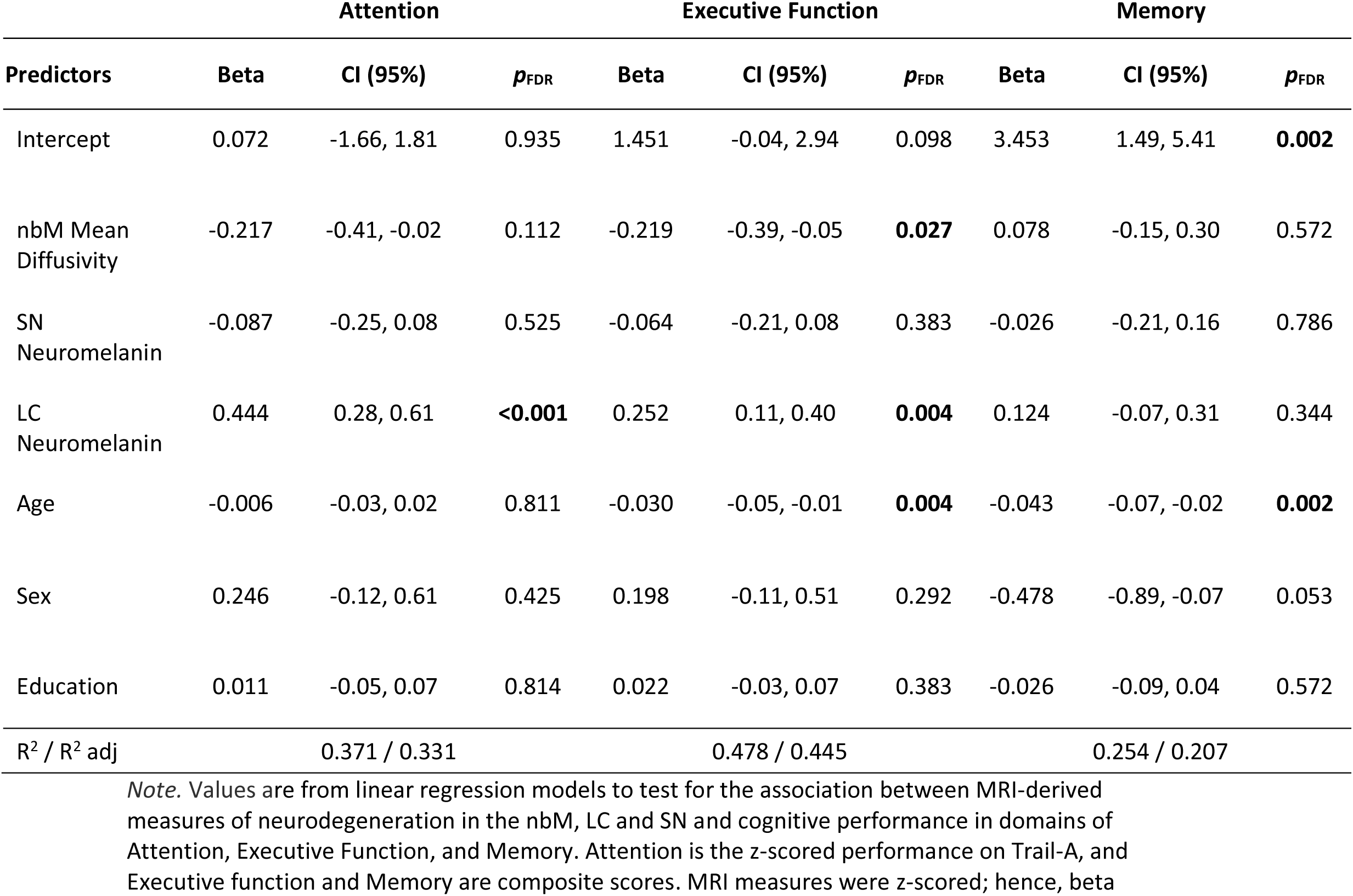
Associations between MRI-derived measures of neurodegeneration and cognitive performance in PD.

### Group differences in the relationship between nbM integrity and cognitive performance

To determine whether the association between nbM MD and cognitive performance differed between groups, we tested for an interaction between nbM MD and group on cognitive performance. For attention, there was no interaction between nbM MD and group (β_nbM*Group_ = - 0.06, 95% CI [−0.34, 0.22], *p*_FDR_ = .684; **Figure 2**). For executive function, there was an interaction, such that the association between higher nbM MD and worse executive function was stronger in PD compared to controls (β_nbM*Group_ = −0.30, 95% CI [−0.54, −0.06], *p*_FDR_ = .036). For memory, there was no interaction between nbM MD and group (β_nbM*Group_ = −0.01, 95% CI [−0.32, 0.29], *p*_FDR_ = .954). Full regression model outputs are presented in **Supplemental Table 5**.

**Figure 2.**
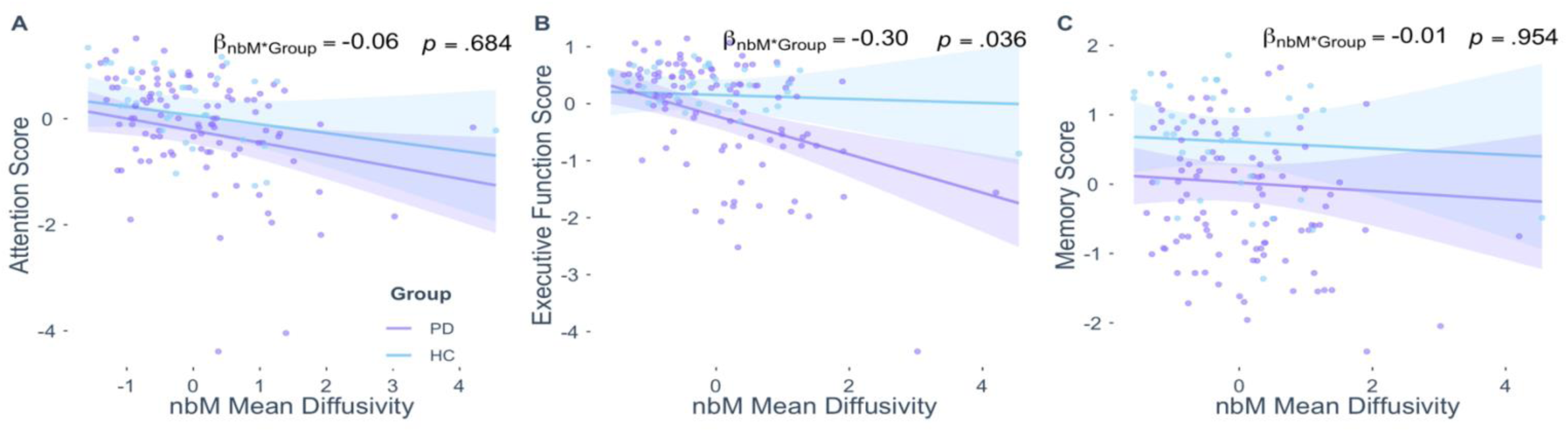
Relationship between nbM degeneration and cognitive performance. (A) Association between nbM MD and Attention score, which is the z-scored performance on Trail-A; (B) association between nbM MD and Executive Function composite score; (C) association between nbM MD and Memory composite score. β estimates reflect the interaction of nbM*Group. *p*-values are FDR-corrected. Linear regression models additionally controlled for LC and SN neuromelanin signal, age, sex, and years of education. Shaded areas reflect 95% confidence intervals.

## DISCUSSION

This study aimed to concurrently measure degeneration in three neurotransmitter systems affected by PD to delineate the unique contribution of nbM degeneration to the cognitive deficits reported in PD. We found that microstructural integrity of the nbM, indexed by DWI-derived MD, did not differ between PD patients and controls. Nevertheless, within PD, lower nbM integrity was independently associated with worse executive function, even after controlling for degeneration of the SN and LC. LC degeneration was also associated with worse executive function. Results were directionally similar when using nbM volume to index degeneration. We also found that the association between nbM integrity and executive function was stronger in PD than in controls, suggesting that loss of nbM integrity exerts a greater impact on executive function in PD than in aging. These findings advance our understanding of the neural and pathological mechanisms underlying early cognitive deficits in PD by suggesting that cholinergic nbM degeneration contributes to executive dysfunction in PD independently of the effects of noradrenergic LC degeneration. However, the relationship may not be exclusively attributable to PD pathology in the earlier stages of the disease.

Several lines of evidence, including post-mortem studies, indicate that PD pathology, namely Lewy bodies and alpha-synuclein fibrils, eventually involves the nbM^14,17,33^. Our results, however, add to the growing body of literature that suggests that during the earlier stages of PD (mean disease duration of 3.9 years in our sample), the degree of microstructural integrity loss observed in PD, as measured by MRI, is no greater than that observed with aging^15,16,34,35^. For instance, recent studies in PD that used volumetric measures of degeneration found that the volumes of the nbM and basal forebrain were similar to those of age-matched healthy controls^16,34,35^. Nevertheless, conflicting findings across studies using volumetric or DWI-derived measures of degeneration and also focus on early-stage PD, before the onset of dementia^14,33^. For instance, one recent study reported greater mean and radial diffusivity of the nbM in PD when relying on histologically-derived segmentations of Ch3 and Ch4^14^, and others have reported smaller nbM volumes in PD patients^33^. One reason for these differences may stem from the definition of the region of interest. Some studies include Ch4 (i.e., the nbM) in addition to other nuclei that make up the basal forebrain^14^, or further segment the Ch4 into anterior and posterior regions, although the relevance of this distinction to the spread of PD pathology is not clear. We chose to focus specifically on the nbM because it has the highest proportion of cholinergic neurons in the brain, in contrast to the Ch3, which contains the lowest proportion of cholinergic neurons, thus providing the largest source of cholinergic projections to cortex^36^. The fact that the pattern of results was consistent across DWI-derived and volumetric measurements also suggests that DWI-derived MD is meaningfully capturing changes to structural integrity. The choice of control group may also explain discrepancies across studies. For instance, some excluded older adults with MoCA scores <27, which may explain why these studies identified group differences whereas we did not^33^. Our findings suggest that when comparing earlier stage PD patients to an unselected group of older adults, nbM microstructural integrity is no lower in early PD than in older adults. This raises questions about the true additive effect of PD neurodegeneration in the nbM. Cholinergic PET studies have shown that cortical cholinergic denervation in early PD is associated with cognitive impairment, but our findings raise the possibility that this relationship may be partly influenced by the concomitant early cortical atrophy known to occur in PD, although an alternate explanation is that terminal loss can be detected earlier than nbM degeneration^7,37^.

Our results demonstrate that nbM degeneration accounts for unique variance in executive function beyond that accounted for by degeneration in the noradrenergic LC. Despite the widely held view that cognitive changes in PD reflect the combined effects of degeneration in multiple ascending neuromodulatory systems, to our knowledge, no previous studies have concurrently examined the nbM and other systems. It is worth noting that we also found a trend for an association between nbM integrity and attention, which was also independent of the association between LC and attention. The nbM provides the primary source of cortical cholinergic innervation, which is critical for the cognitive control required for performance on executive function tasks^38^, whereas the LC modulates arousal and cognitive flexibility, which are also required of executive function tasks^39^. Our results therefore suggest that early disruption in these parallel systems may act additively, through somewhat different mechanisms, to impair executive function. Indeed, it is interesting to note that the effect size of the nbM association with executive function was comparable to that of the LC. This has important clinical implications as it indicates that interventions targeting cholinergic dysfunction may be beneficial even in the presence of LC degeneration, and vice versa.

Finally, although the association between the nbM integrity and executive function was stronger in PD, the data do not establish that it is exclusive to PD. This finding aligns with a large body of research showing a relationship between nbM integrity (and the basal forebrain, more generally) and cognitive function in aging and in Alzheimer’s disease^8,12,13^. This has important research implications, as it suggests that MRI-derived measures of nbM degeneration may not be suitable as biomarkers of cognitive impairment if the goal is to track PD-specific changes to the nbM. Nonetheless, as the relationship between nbM microstructural integrity and executive function was stronger in PD than in controls, measuring the nbM may provide insights into a possible loss of compensatory mechanisms, as might be caused, for instance, by early PD-associated cortical atrophy^37^.

An important limitation of our study is that we did not have access to biomarker measurements to quantify Alzheimer’s disease-associated degeneration, which would have allowed us to determine whether the similarities between PD patients and controls are driven by a comparable presence of AD pathology^40^. Determining whether nbM degeneration is causally related to cognitive decline will require longitudinal investigation. Future work would also benefit from measuring the integrity of the white matter projections from the nbM to cortical regions known to support attention, executive function, and memory^37^.

To summarize, our findings support the notion that loss of integrity in the cholinergic nbM contributes to executive dysfunction in PD patients. This effect is independent of LC degeneration, suggesting cholinergic and noradrenergic dysfunction may represent partly separable therapeutic targets for remediating cognitive dysfunction. Our study also highlights the importance of jointly considering the effects of different aspects of neurodegeneration when aiming to identify neural correlates of the clinical phenotype of PD.

## Data Availability

All data produced in the present study are available upon reasonable request to the authors.

## Acknowledgements

We thank Dr. Alexander Wiesman, Ms. Rebekah Wickens, Dr. Jason da Silva Castenheira, and Dr. R. Nathan Spreng for helpful discussions. We also thank the individuals with PD and the controls for their participation, and the Quebec Parkinson Network for supporting the registry and assisting with participant recruitment.

## Author contributions

AB: conception, analysis, writing, and revising the manuscript.

ESO: design and revising the manuscript.

VM: data acquisition, analysis, and revising the manuscript.

NS: analysis and revising the manuscript.

JFG: design, analysis, and revising the manuscript.

AD: funding and revising the manuscript.

DLC: analysis, revising the manuscript.

MS: conception, design, funding, supervision, writing, and revising the manuscript.

