## Supplementary_Tables_Figures for "Nucleus basalis of Meynert integrity is associated with cognitive dysfunction in Parkinson’s disease independently of locus coeruleus degeneration"

### SUPPLEMENTARY FIGURES AND TABLES

#### Supplemental Table 1. T1 MRI sequence parameters

| MRI sequence name | Dimension | Relaxation time, TR | Echo time, TE | Flip Angle | Field of view for 2D acquisitions | Slice thickness | 3D volumetric resolution | Slice orientation | Matrix size (xyz) | Voxel size |
|---|---|---|---|---|---|---|---|---|---|---|
| 3D T1 Magnetization Prepared – Rapid Gradient Echo (MPRAGE) | 3D | 2300.0 ms | 2.98 ms | 9° | NA | 1.0 mm | 192 x 256 x 256 $mm^3$ | Sagittal | 192 x 256 x 256 | 1 x 1 x 1 $mm^3$ |
| T1-weighted Fast Spin Echo (neuromelanin-MRI) | 2D | 600.0 ms | 10 ms | 120° | 165 x 220 $mm^2$ | 1.8 mm | NA | T > C4.9 | 240 x 320 x 20 | 0.7 x 0.7 x 1.8 $mm^3$ |

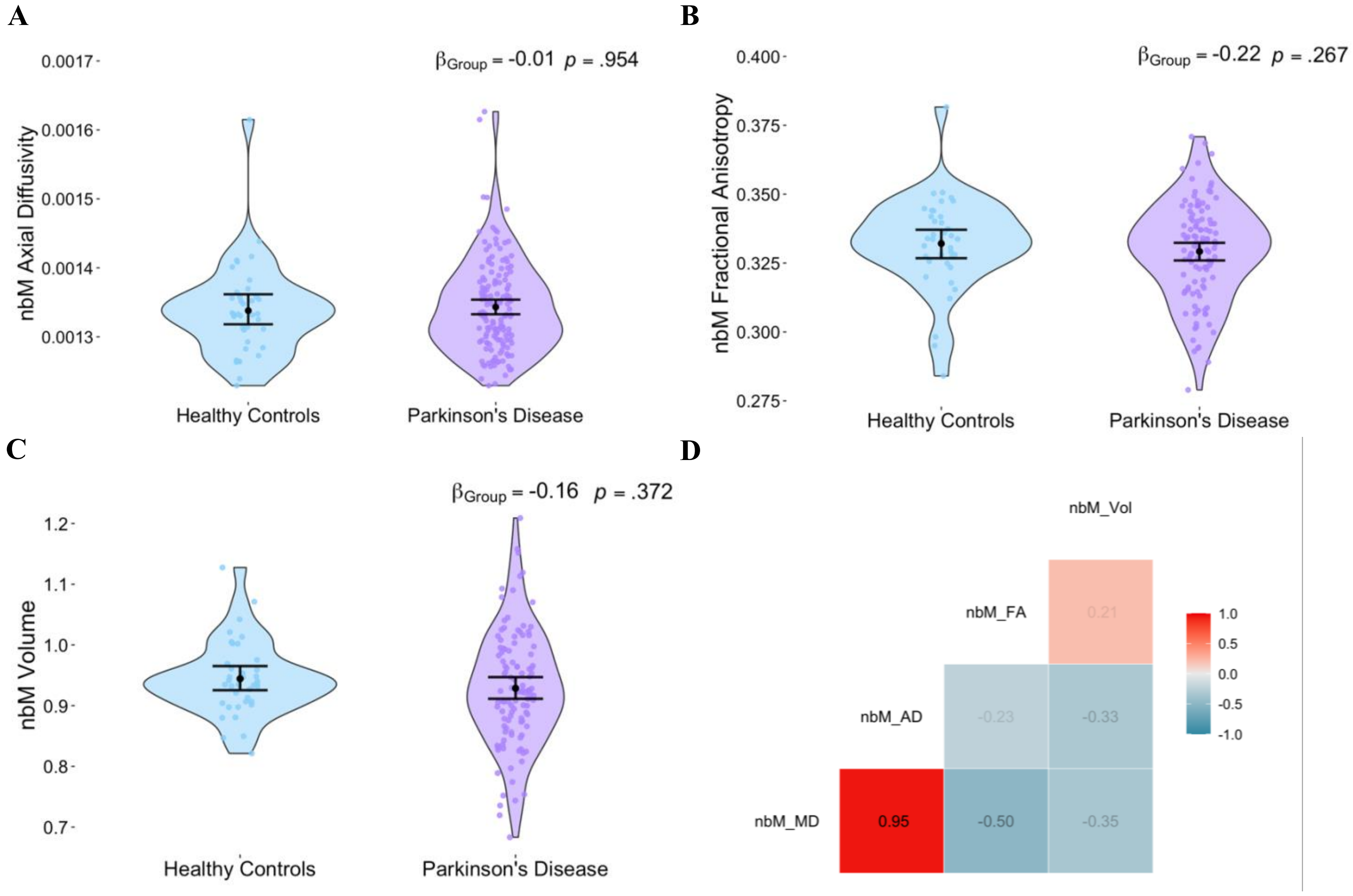


**Supplemental Figure 1**. **Group difference in MRI-derived measures of nbM degeneration.** A) Violin plot depicting axial diffusivity (AD); B) fractional anisotropy (FA); and C) DBM-derived volume (average of right and left) in Controls and Parkinson's patients, where values represent deviation from a template; D) Correlation matrix showing pair-wise correlations of the MRI-derived measures of nbM degeneration in PD patients. Beta estimates and *p*-values for group differences were extracted from linear regression models controlling for age, sex, and years of education. Error bars reflect 95% bootstrapped confidence intervals.

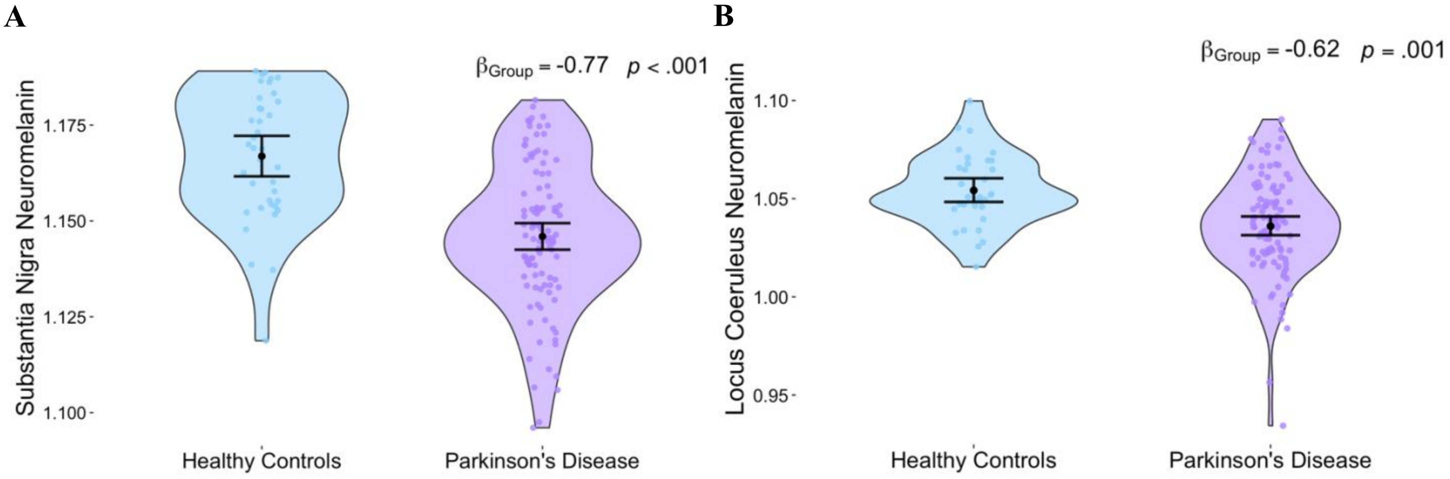


**Supplemental Figure 2**. **Group differences in MRI-derived neuromelanin signal.** A) substantia nigra and B) locus coeruleus. Violin plots of mean diffusivity of the nucleus basalis of Meynert (average of right and left) in Controls and Parkinson's patients. Beta estimates and p-values for group differences were obtained from linear regressions that controlled for age, sex, and years of education. Error bars reflect 95% bootstrapped confidence intervals.

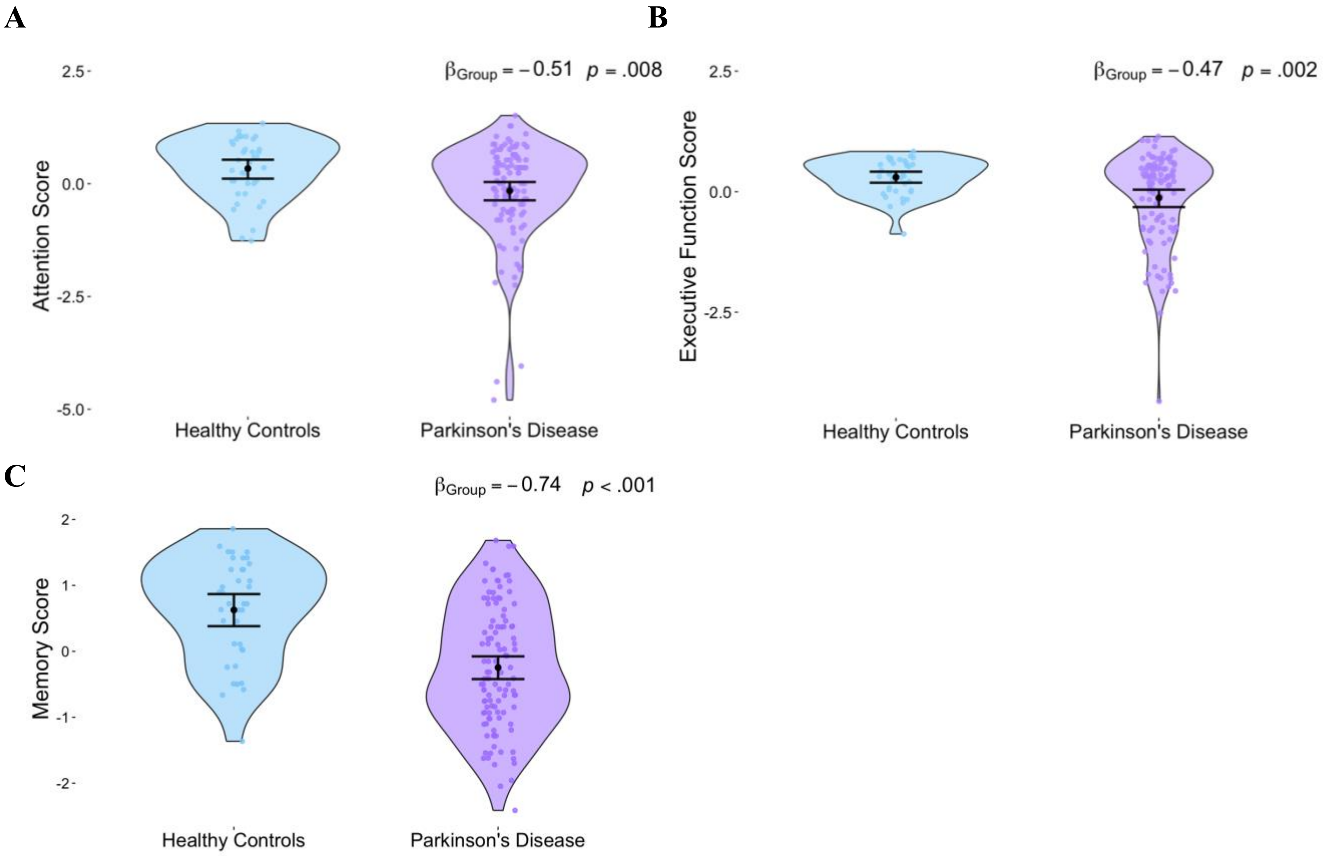


**Supplemental Figure 3**. **Group differences in cognitive performance.** A) attention, B) executive function, and C) memory. Estimates of group differences (Controls and Parkinson's patients) obtained from linear regressions on each cognitive domain that controlled for age, sex and years of education. Error bars reflect 95% bootstrapped confidence intervals.

**Supplemental Table 2. Performance on neuropsychological tests**

| Data | n | Healthy Controls | n | Parkinson's Disease | *p* value |
|---|---|---|---|---|---|
| ***Attention Measure*** | | | | | |
| Trail Making Test A | 46 | 32.63 (11.33) | 112 | 41.81 (18.59) | **<.001** |
| ***Executive Function Measures*** | | | | | |
| Trail Making Test B-A | 46 | 38.89 (24.07) | 108 | 63.84 (59.59) | **<.001** |
| Brixton Error | 46 | 16.67 (5.68) | 110 | 18.64 (8.63) | 0.097 |
| Executive Function Composite Score | 46 | 0.22 (0.4) | 112 | -0.1 (0.82) | **0.001** |
| ***Memory Measures*** | | | | | |
| HVLT-R Total Score Trials I, II, III | 46 | 26.33 (4.41) | 112 | 21.49 (5.52) | **<.001** |
| HVLT-R 4 | 46 | 9.61 (2.37) | 112 | 7.2 (2.86) | **<.001** |
| Memory Composite Score | 46 | 0.59 (0.76) | 112 | -0.24 (0.92) | **<.001** |

*Note.* Significant *p*-values are listed in bold, all comparisons conducted using Welch's t-tests. HVLT-R, Hopkins Verbal Learning Test–Revised.

**Supplemental Table 3. Associations between MRI-derived measures of neurodegeneration and cognitive performance in PD, without multiple comparisons**

| | Attention | | | Executive Function | | | Memory | | |
|---|---|---|---|---|---|---|---|---|---|
| *Predictors* | *Beta* | *CI (95%)* | *p* | *Beta* | *CI (95%)* | *p* | *Beta* | *CI (95%)* | *p* |
| Intercept | 0.072 | -1.66, 1.81 | 0.935 | 1.451 | -0.04, 2.94 | 0.056 | 3.453 | 1.49, 5.41 | **0.001** |
| Mean Diffusivity nbM | -0.217 | -0.41, -0.02 | **0.032** | -0.219 | -0.39, -0.05 | **0.012** | 0.078 | -0.15, 0.30 | 0.491 |
| Neuromelanin SN | -0.087 | -0.25, 0.08 | 0.300 | -0.064 | -0.21, 0.08 | 0.376 | -0.026 | -0.21, 0.16 | 0.786 |
| Neuromelanin LC | 0.444 | 0.28, 0.61 | **<0.001** | 0.252 | 0.11, 0.40 | **0.001** | 0.124 | -0.07, 0.31 | 0.197 |
| Age | -0.006 | -0.03, 0.02 | 0.579 | -0.030 | -0.05, -0.01 | **0.001** | -0.043 | -0.07, -0.02 | **0.001** |
| Sex | 0.246 | -0.12, 0.61 | 0.182 | 0.198 | -0.11, 0.51 | 0.209 | -0.478 | -0.89, -0.07 | **0.023** |
| Education (Years) | 0.011 | -0.05, 0.07 | 0.698 | 0.022 | -0.03, 0.07 | 0.383 | -0.026 | -0.09, 0.04 | 0.441 |
| $R^2$ / $R^2$ adjusted | 0.371 / 0.331 | | | 0.478 / 0.445 | | | 0.254 / 0.207 | | |

*Note.* Values are from linear regression models to test for the association between MRI-derived measures of neurodegeneration in the nbM, LC and SN and cognitive performance in domains of Attention, Executive Function, and Memory. Attention is the z-scored performance on Trail-A, and Executive function and Memory are composite scores. MRI measures were z-scored; hence, beta estimates represent the change in cognitive domain score associated with a 1 SD change in the MRI measure. Sex was coded as 0 = Female and 1 = Male, Education was measured in years. *p*-values are non-FDR–adjusted *p*-values, and values < 0.05 are bolded.

**Supplemental Table 4. Association between different MRI-derived measures of nbM degeneration and cognitive performance in PD**

**A) Association between nbM axial diffusivity and cognition in PD**

| | | Attention | | | Executive Function | | | Memory | |
|---|---|---|---|---|---|---|---|---|---|
| *Predictors* | *Beta* | *CI (95%)* | *p*(FDR) | *Beta* | *CI (95%)* | *p*(FDR) | *Beta* | *CI (95%)* | *p*(FDR) |
| Intercept | 0.353 | -1.48, 2.19 | 0.769 | 2.117 | 0.48, 3.76 | 0.028 | 3.130 | 1.27, 4.99 | 0.004 |
| AD nbM | -0.166 | -0.38, 0.04 | 0.408 | -0.132 | -0.32, 0.05 | 0.283 | 0.077 | -0.13, 0.29 | 0.547 |
| SN Neuromelanin | -0.116 | -0.31, 0.08 | 0.421 | -0.087 | -0.26, 0.08 | 0.393 | -0.019 | -0.22, 0.18 | 0.847 |
| LC Neuromelanin | 0.480 | 0.30, 0.66 | **<0.001** | 0.281 | 0.12, 0.44 | **0.003** | 0.117 | -0.07, 0.30 | 0.363 |
| Age | -0.011 | -0.03, 0.01 | 0.458 | -0.041 | -0.06, -0.02 | **0.001** | -0.041 | -0.06, -0.02 | **0.003** |
| Sex | 0.238 | -0.16, 0.63 | 0.421 | 0.171 | -0.18, 0.52 | 0.393 | -0.465 | -0.86, -0.07 | 0.053 |
| Education | 0.009 | -0.05, 0.07 | 0.769 | 0.019 | -0.04, 0.08 | 0.502 | -0.025 | -0.09, 0.04 | 0.547 |
| $R^2$ / $R^2$ adjusted | 0.356 / 0.315 | | | 0.451 / 0.416 | | | 0.255 / 0.208 | | |

*Note.* Linear regression models to test for the association between nbM axial diffusivity (AD) as a proxy measure of microstructural integrity in the nbM and cognitive performance in domains Attention, Executive Function, and Memory. MRI measures were z-scored; hence, beta estimates represent the change in cognitive domain score associated with a 1 SD change in the MRI measure. Attention is the z-scored performance on Trails A, and Executive function and Memory are composite scores. Sex was coded as 0 = Female and 1 = Male, Education was measured in years. *p*-values are the FDR–adjusted *p*-values, and values < 0.05 are bolded.

**B) Association between nbM fractional anisotropy and cognition in PD**

| | Attention | | | Executive Function | | | Memory | | |
|---|---|---|---|---|---|---|---|---|---|
| *Predictors* | *Beta* | *CI (95%)* | *p*(FDR) | *Beta* | *CI (95%)* | *p*(FDR) | *Beta* | *CI (95%)* | *p*(FDR) |
| Intercept | 0.786 | -0.84, 2.41 | 0.477 | 2.086 | 0.70, 3.48 | **0.006** | 2.831 | 1.18, 4.48 | **0.003** |
| FA nbM | 0.128 | -0.05, 0.30 | 0.338 | 0.241 | 0.09, 0.39 | **0.004** | -0.023 | -0.20, 0.15 | 0.795 |
| SN Neuromelanin | -0.054 | -0.26, 0.15 | 0.694 | 0.008 | -0.17, 0.18 | 0.925 | -0.036 | -0.24, 0.17 | 0.795 |
| LC Neuromelanin | 0.467 | 0.29, 0.65 | **<0.001** | 0.261 | 0.11, 0.42 | **0.004** | 0.120 | -0.06, 0.30 | 0.345 |
| Age | -0.018 | -0.04, 0.001 | 0.245 | -0.042 | -0.06, -0.03 | **<0.001** | -0.037 | -0.06, -0.02 | **0.002** |
| Sex | 0.209 | -0.18, 0.60 | 0.477 | 0.171 | -0.16, 0.50 | 0.415 | -0.445 | -0.84, -0.05 | 0.064 |
| Education | 0.010 | -0.05, 0.07 | 0.754 | 0.025 | -0.03, 0.08 | 0.415 | -0.023 | -0.09, 0.04 | 0.658 |
| $R^2$ / $R^2$ adj | 0.354 / 0.313 | | | 0.496 / 0.464 | | | 0.251 / 0.204 | | |

*Note.* Linear regression models to test for the association between nbM fractional anisotropy (FA) as a proxy measure of microstructural integrity and cognitive performance in domains of Attention, Executive Function, and Memory. MRI measures were z-scored; hence, beta estimates represent the change in cognitive domain score associated with a 1 SD change in the MRI measure. Attention is the z-scored performance on Trails A, and Executive function and Memory are composite scores. Sex was coded as 0 = Female and 1 = Male, Education was measured in years. *p*-values are the FDR–adjusted *p*-values, and values < 0.05 are bolded.

**C) Association between nbM volume and cognition in PD**

| | Attention | | | Executive Function | | | Memory | | |
|---|---|---|---|---|---|---|---|---|---|
| *Predictors* | *Beta* | *CI (95%)* | *p*(FDR) | *Beta* | *CI (95%)* | *p*(FDR) | *Beta* | *CI (95%)* | *p*(FDR) |
| Intercept | 0.525 | -1.20, 2.25 | 0.637 | 1.969 | 0.46, 3.48 | **0.026** | 1.776 | 0.10, 3.46 | 0.068 |
| nbM Volume | 0.142 | -0.03, 0.31 | 0.372 | 0.179 | 0.03, 0.33 | **0.036** | 0.233 | 0.07, 0.40 | **0.050** |
| SN Neuromelanin | -0.116 | -0.31, 0.08 | 0.418 | -0.097 | -0.27, 0.07 | 0.369 | -0.060 | -0.25, 0.13 | 0.624 |
| LC Neuromelanin | 0.468 | 0.29, 0.65 | **<0.001** | 0.267 | 0.11, 0.43 | **0.004** | 0.105 | -0.07 – 0.28 | 0.338 |
| Age | -0.014 | -0.04, 0.01 | 0.400 | -0.040 | -0.06, -0.02 | **<0.001** | -0.024 | -0.04, -0.004 | 0.068 |
| Sex | 0.200 | -0.19, 0.59 | 0.434 | 0.145 | -0.20, 0.49 | 0.399 | -0.423 | -0.80, -0.04 | 0.068 |
| Education | 0.012 | -0.05, 0.08 | 0.715 | 0.024 | -0.03, 0.08 | 0.399 | -0.011 | -0.07, 0.05 | 0.721 |
| $R^2$ / $R^2$ adjusted | 0.357 / 0.317 | | | 0.471 / 0.437 | | | 0.306 / 0.262 | | |

*Note.* Linear regression models to test for the association between nbM volume as a proxy measure of microstructural integrity and cognitive performance in domains of Attention, Executive Function, and Memory. MRI measures were z-scored; hence, beta estimates represent the change in cognitive domain score associated with a 1 SD change in the MRI measure. Attention is the z-scored performance on Trails A, and Executive function and Memory are composite scores. Sex was coded as 0 = Female and 1 = Male, Education was measured in years. *p*-values are the FDR–adjusted p-values, and values < 0.05 are bolded.

**Supplemental Table 5. Group differences in the association between nbM mean diffusivity and cognition**

| | Attention | | | EF | | | Memory | | |
|---|---|---|---|---|---|---|---|---|---|
| *Predictors* | *Beta* | *CI (95%)* | *p*(FDR) | *Beta* | *CI (95%)* | *p*(FDR) | *Beta* | *CI (95%)* | *p*(FDR) |
| Intercept | 0.596 | -0.68, 1.88 | 0.525 | 1.189 | 0.10, 2.28 | 0.059 | 1.540 | 0.16, 2.920 | 0.091 |
| nbM Mean Diffusivity | -0.166 | -0.42, 0.09 | 0.389 | -0.035 | -0.25, 0.18 | 0.750 | -0.046 | -0.32, 0.23 | 0.952 |
| SN Neuromelanin | -0.065 | -0.22, 0.09 | 0.525 | -0.059 | -0.19, 0.07 | 0.460 | -0.005 | -0.17, 0.16 | 0.954 |
| LC Neuromelanin | 0.411 | 0.26, 0.56 | **<0.001** | 0.264 | 0.14, 0.39 | **0.001** | 0.172 | 0.01, 0.34 | 0.091 |
| Age | -0.010 | -0.03, 0.01 | 0.389 | -0.020 | -0.03, -0.01 | **0.026** | -0.020 | -0.04, -0.002 | 0.091 |
| Sex | 0.246 | -0.07, 0.56 | 0.370 | 0.222 | -0.05, 0.49 | 0.154 | -0.272 | -0.61, 0.07 | 0.206 |
| Education | 0.009 | -0.04, 0.05 | 0.684 | 0.016 | -0.02, 0.05 | 0.460 | 0.021 | -0.03, 0.07 | 0.563 |
| Group | -0.285 | -0.64, 0.07 | 0.370 | -0.369 | -0.67, -0.07 | **0.036** | -0.587 | -0.97, -0.21 | **0.024** |
| Interaction nbM*Group | -0.061 | -0.34, 0.22 | 0.684 | -0.302 | -0.54, -0.06 | **0.036** | -0.014 | -0.32, 0.29 | 0.954 |
| $R^2$ / $R^2$ adjusted | 0.370 / 0.331 | | | 0.459 / 0.426 | | | 0.305 / 0.261 | | |

*Note.* Linear regression models to test for an interaction between nbM mean diffusivity and Group on cognitive performance in domains of Attention, Executive Function, and Memory. MRI measures were z-scored; hence, beta estimates represent the change in cognitive domain score associated with a 1 SD change in the MRI measure. Attention is the z-scored performance on Trails A, and Executive function and Memory are composite scores. Group was coded as 0 = HC; 1 = PD. Sex was coded as 0 = Female and 1 = Male, Education was measured in years. *p*-values < 0.05 after FDR correction are bolded.
